# Evaluation of User Experiences for the Clean Team Ghana Container-Based Sanitation Service in Kumasi, Ghana

**DOI:** 10.1101/2020.10.23.20218578

**Authors:** James B. Tidwell, Kwabena B. Nyarko, Ian Ross, Bismark Dwumfour-Asare, Pippa Scott

## Abstract

There is a lack of affordable and acceptable sanitation solutions for dense, low-income urban settlements. One option that has been proposed is container-based sanitation, where a sealed cartridge installed in a free-standing toilet is regularly changed and adequately separates users from their excreta. Though container-based toilets are considered a safely managed sanitation solution that meets the Sustainable Development Goal for sanitation, little is known about user experiences to inform how such a solution should be viewed by governments. We conducted a longitudinal prospective cohort study of changes in objective and subjective measures of sanitation quality due to the Clean Team Ghana (CTG) container-based toilet service in Kumasi, Ghana from June to December 2019. We collected data immediately prior to installation of a toilet and 10 weeks afterwards for 292 customers. Most initially used public toilets with good structural quality, but sometimes had poor cleanliness, a lack of handwashing facilities, and required a 14.3 minute mean round trip time to use. The evaluation found that CTG delivers a high-quality service that positively impacts the quality of life of CTG customers, as well as saving them money, reducing gender gaps in quality of life, and addressing the needs of those with physical disabilities. Satisfaction with CTG toilet and service characteristics was high, with the largest increases for satisfaction with smell, comfort, disgust, and privacy. Women in particular were positively impacted both for explicitly gendered indicators like the ability to practice menstrual hygiene management, and other indicators where women scored lower than men at baseline, including ease of access, ease of use, and cleanliness. Use of the service also benefitted those who had been unable to use a toilet for physical or social reasons beforehand. Problems with the CTG service, such as leaking, filling, smelling, or not being replaced as scheduled, were reported by fewer than 10% of customers. While one product or service does not fit the needs of all customers, this evaluation supports the growing body of evidence that container-based sanitation provides a service valued by users and acceptable to policymakers in dense urban settlements.

## Introduction

About 4.2 billion people lack access to safely managed sanitation globally (World Health Organization, 2019) with more than half residing in urban areas, where shared sanitation is more common than in rural areas. The high density and rapidly expanding fringes of cities and towns regularly present service delivery challenges for local and national governments in low- and middle-income countries (Ezeh et al., 2017). As urban populations grow, services should expand to meet their needs, but networked sanitation is not extending nearly rapidly enough to provide the necessary services (Öberg, Metson, Kuwayama, & A Conrad, 2020). On-site sanitation systems such as improved pit latrines and septic tanks become less appropriate as population density increases due to the safe management requirements of pit leachate, septage effluent and the safe removal of solids (Amin et al., 2020). The concept of Citywide Inclusive Sanitation (CWIS) seeks to overcome this technology dichotomy by focusing on service outcomes and promoting a range of solutions—both onsite and sewered, centralized or decentralized – to ensure everyone has equitable access to safely managed sanitation (Schrecongost, Pedi, Rosenboom, Shrestha, & Ban, 2020).

Container Based Sanitation (CBS) is a system where sealed toilet containers are regularly collected from households and transported to treatment facilities. CBS, if properly managed, ensures full containment of faecal waste, by contrast with onsite-infiltration systems (latrines, septic tanks) which usually discharge contaminated liquid effluent locally (Peal et al., 2020). Several CBS business models have emerged globally since 2010 providing a viable, low-cost sanitation option. They are particularly relevant for low-income urban settlements where there is relatively higher ability to pay for sanitation services and on-site sanitation and sewerage would not be technically feasible or cost-efficient (Nyoka et al., 2017; O’Keefe, Luthi, Tumwebaze, & Tobias, 2015; K. Russel et al., 2015; Tilmans et al., 2015). Studies of CBS have primarily focused on business models, costs, potential health impacts, and environmental exposures for users and service providers. Findings include that services are generally considered safe by users (World Bank, 2019), often ensure fecal pathogens are killed before disposal (Preneta, Kramer, Magloire, & Noel, 2013), and are more resilient than some other kinds of sanitation services to floods and droughts. They also disproportionately benefit the elderly and young (Greenland et al., 2016), facilitate handwashing with soap (Greenland et al., 2016), and are a cost-efficient way to expand services to marginalized communities (World Bank, 2019). However, CBS businesses rarely cover their costs through business revenues (K. C. Russel et al., 2019).

Given this, understanding user experiences may lead to opportunities for service improvements and improved marketing approaches to leverage existing willingness to pay (Tidwell, Terris-Prestholt, Quaife, & Aunger, 2019) and to increase acceptability to governments. Additionally, understanding impacts on vulnerable groups and the potential market for inclusive solutions are essential for ensuring better and more equitable sanitation service delivery.

In Kumasi, Ghana, 36% of the population use public toilets, 11% use pit latrines, 7% use Kumasi Ventilated Improved Pit (KVIP) latrines (Ghana Statistical Service, 2010). Only 2% use no toilet facilities, a low figure due to both high population density and the ubiquity of public toilets. The remainder (43%), primarily concentrated in wealthier areas use private household flushing toilets. There are some ongoing initiatives for promoting fixed toilets, including Water and Sanitation for the Urban Poor (WSUP)’s “Toilet in Every Compound” program, which used market-based incentives resulting in 405 new household toilets in 2019 (USAID, PSI, PATH, & WSUP, 2019). The Government of Ghana through the Kumasi Metropolitan Assembly is also providing 400 household toilet facilities to some poor households in Kumasi in 2020 (Bonsu, 2020). Clean Team Ghana (CTG) introduced a CBS solution in Kumasi in 2010 and by 2020 are serving more than 2,800 low-income residents. The primary aim of this study was to understand user experience of the CTG service from both quality of service and quality of life perspectives. Underling objectives included characterizing the CTG user base, evaluating CTG service delivery quality, examining gender differences in the user experience of CTG service, and exploring cost implications for users compared to other sanitation options.

## Methods

### STUDY SETTING

This study surveyed customers of a container-based sanitation (CBS) service provider operating in Kumasi, Ghana. Clean Team Ghana (CTG) is a social enterprise delivering CBS services comprising a high-quality plastic toilet with a sealable internal waste container. CTG operates on a subscription model: customers pay a monthly fee for weekly collection and replacement of the waste container, and the toilet remains property of CTG. At the start of the study (June 2019), the business was serving about 1,500 customers and entering a phase of scale-up. The CTG model in Kumasi has been refined over the past 10 years and is now relatively stable. The primary alternative open to Kumasi’s low-income population are 400 independently-operated public toilets, most originally constructed by the government, providing a fairly homogenous comparator service from which most new customers are likely to be transitioning (Container Based Sanitation Alliance, 2018). The pay-per-use public toilet model, from which CBS is likely an upgrade in terms of offering a private, household toilet, is very common in Ghana, but is also present in many other countries. As the second-largest city in Ghana with a third of its 2.7 million population using public toilets, Kumasi represents a potentially large market for future expansion. Kumasi was therefore an appropriate setting to undertake research to explore user experiences of CBS for the sector.

### STUDY DESIGN

We conducted a longitudinal prospective cohort study of new CTG customers. Respondents were surveyed twice – shortly before the installation of the CTG toilet and then 10 weeks after its installation. These time points were selected to assess experiences after some period of adapting to the benefits and challenges of using a new technology. The purpose of the evaluation was to understand the impact on self-selected customers, so no control group was used as a comparison and no random allocation of households to kinds of sanitation was performed.

We estimated that a sample size of 360 customers would be enrolled at baseline, with an assumed loss to follow up of 5% to allow for a minimum detectable change of 7.4% with a power of 80% and a 95% confidence.

The outcomes assessed include measures of service quality (e.g., product failures, frequency and regularity of pick-ups), objective measures of sanitation quality (e.g., cleanliness, accessibility, and other measures from the Healthy Sanitation Framework (Tidwell, Chipungu, Chilengi, & Aunger, 2018)), and subjective measures of service satisfaction (e.g. with comfort). In addition, users’ Sanitation-related Quality of Life (SanQoL) was assessed across five attributes (disgust, health, privacy, shame, safety).

### DATA COLLECTION

Quantitative household surveys started in June 2019 following two weeks of enumerators’ training and piloting of tools and protocols. We adopted an electronic data collection approach using mobile phones KoBoToolbox software. This provided time- and location- stamped data and photographs of sanitation conditions and CTG installations for accountability purposes. Key terminology and text items in the questionnaire were translated by Akan Twi language experts from the Department of Languages of the Kwame Nkrumah University of Science and Technology (Kumasi, Ghana) to adequately reflect the concepts in the English versions. We also undertook cognitive interviews focused on SanQoL items, to ensure the translations adequately captured key concepts, and that the five attributes were relevant and acceptable in the setting. The surveys captured information on sanitation service levels, quality and practices. Baseline surveys were administered to new CTG customers after they expressed a desire to enrol, but prior to the installation of the container-based toilet facility, to reduce risk of bias due to recall or a new perspective on the prior service. Enumerators directly observed sanitation conditions at both baseline and endline.

### ANALYSIS

Basic descriptive statistics were calculated to profile customers’ demographic characteristics, tenancy status, prior sanitation quality, and living conditions, as well as the quality of the CTG service. Mean scores for satisfaction and SanQoL were calculated and compared before and after CTG service enrolment using a linear regression model estimated using ordinary least squares. Sub-group analysis by gender used the same approach, but estimated difference-in-differences to understand differential impacts of the service. The “living environment” of CTG customers was assessed using the Living Conditions Diamond tool (LCD) (Gulyani & Bassett, 2010). This objectively characterises different types of settlements in terms of tenure, infrastructure, housing, and neighbourhood attributes (Scott, Cotton, & Sohail, 2015).

### ETHICAL CONSIDERATIONS

Customers were informed about the study before having their new CTG toilet installed. They were told that they would receive no direct benefits from participation in the study, and that they could refuse to participate and still enrol in the service with no negative consequences. Names of respondents were recorded along with coded ID numbers on a separate sheet from the data entry to ensure the same individuals responded at endline, while protecting their anonymity by withholding their name from the dataset. No analysis was conducted at a level where personally identifying information was revealed, and no names or identifiers are used in any outputs of this study. The research protocol was approved by the Committee on Human Research, Publications and Ethics, Kwame Nkrumah University of Science and Technology (KNUST), School of Medical Sciences & Komfo Anokye Teaching Hospital on 22 May 2019 (ref: CHRPE/AP/317/19). This study posed a minimal risk to participants. Survey questions did not address sensitive topics in most cases. Some questions on gendered aspects of sanitation and past experiences were included that may have brought up troubling memories, but there were no questions probing deeply into fears or traumatic experiences of participants.

## Results

We enrolled 404 customers at baseline and completed 292 surveys at endline. The overall attrition rate of 28% consisted of customer dropout from CTG service subscription (17%), unavailability of active customers for the survey (2%), and potential customers subsequently declining to install the toilet (9%). The reasons for customer dropout after service subscription are shown in Supplementary Table 1.

### CUSTOMER CHARACTERISTICS

A majority of customers surveyed at both baseline and endline were female (73%), with a mean age of 44 years (Table 1). There was almost an even split between customers who owned their current residence (45%) and those who rented (52%). Amongst renters, a majority had no landlord present on the plot (60%). Mean rent for tenants was 67 Ghanaian cedi (GHS) per month (about US$ 11.80). Most homes had either one (71%) or two (23%) rooms. Mean duration of living in their current housing was relatively long both amongst owners (17.3 years) and renters (6.5 years). Mean household size was 3.2 people, but many customers had only one (13%) or two (26%) household members, and only 12% had more than five. Respondents lived in dwellings with one (71%) or two (23%) rooms on plots with a mean of 6 other households, which were typically several single-story buildings divided up into separate residences. Most respondents (67%) did not have a water source in their dwelling or on their yard/plot.

**Table 1:**
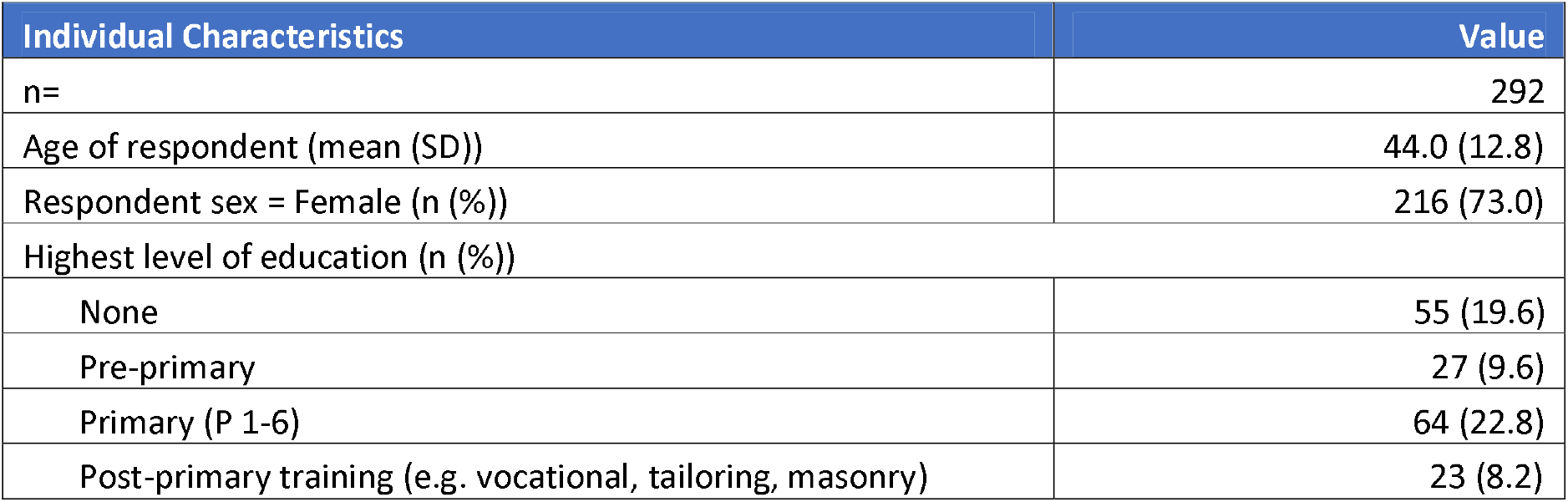
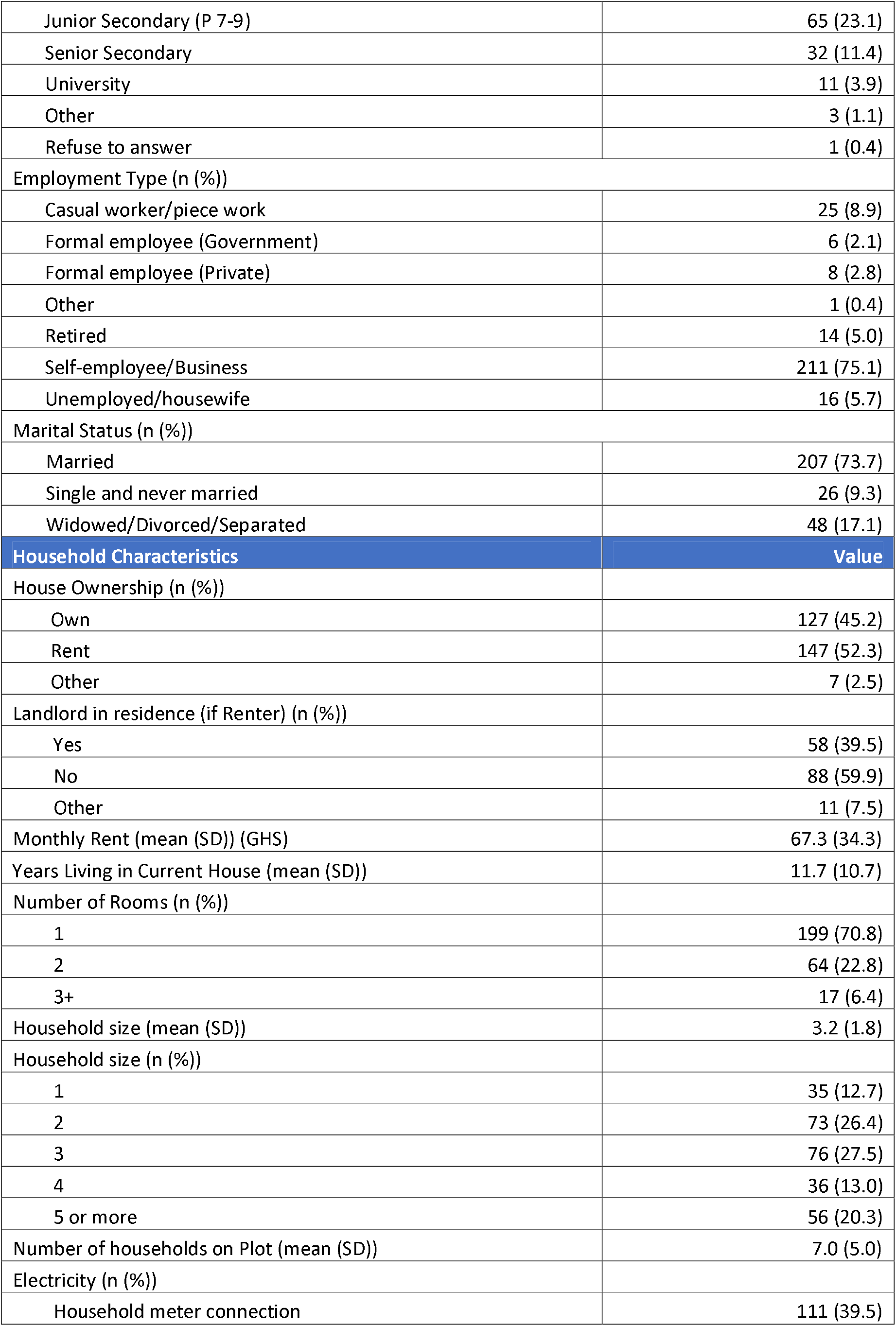
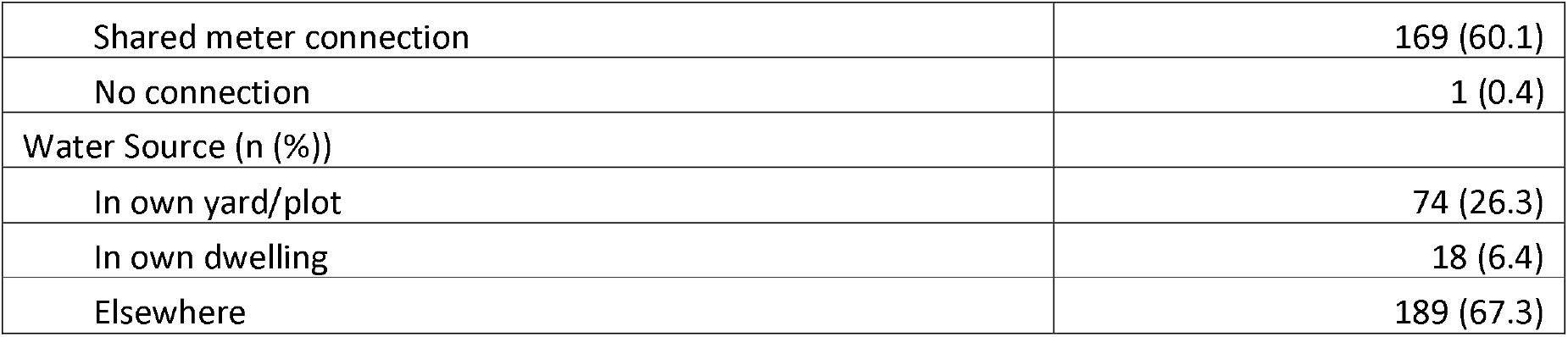
Demographic and Household Characteristics for Clean Team Ghana Customers

| Individual Characteristics | Value |
| --- | --- |
| n= | 292 |
| Age of respondent (mean (SD)) | 44.0 (12.8) |
| Respondent sex = Female (n (%)) | 216 (73.0) |
| Highest level of education (n (%)) |  |
| None | 55 (19.6) |
| Pre-primary | 27 (9.6) |
| Primary (P 1-6) | 64 (22.8) |
| Post-primary training (e.g. vocational, tailoring, masonry) | 23 (8.2) |
| Junior Secondary (P 7-9) | 65 (23.1) |
| Senior Secondary | 32 (11.4) |
| University | 11 (3.9) |
| Other | 3 (1.1) |
| Refuse to answer | 1 (0.4) |
| Employment Type (n (%)) |  |
| Casual worker/piece work | 25 (8.9) |
| Formal employee (Government) | 6 (2.1) |
| Formal employee (Private) | 8 (2.8) |
| Other | 1 (0.4) |
| Retired | 14 (5.0) |
| Self-employee/Business | 211 (75.1) |
| Unemployed/housewife | 16 (5.7) |
| Marital Status (n (%)) |  |
| Married | 207 (73.7) |
| Single and never married | 26 (9.3) |
| Widowed/Divorced/Separated | 48 (17.1) |
| **Household Characteristics** | **Value** |
| House Ownership (n (%)) |  |
| Own | 127 (45.2) |
| Rent | 147 (52.3) |
| Other | 7 (2.5) |
| Landlord in residence (if Renter) (n (%)) |  |
| Yes | 58 (39.5) |
| No | 88 (59.9) |
| Other | 11 (7.5) |
| Monthly Rent (mean (SD)) (GHS) | 67.3 (34.3) |
| Years Living in Current House (mean (SD)) | 11.7 (10.7) |
| Number of Rooms (n (%)) |  |
| 1 | 199 (70.8) |
| 2 | 64 (22.8) |
| 3+ | 17 (6.4) |
| Household size (mean (SD)) | 3.2 (1.8) |
| Household size (n (%)) |  |
| 1 | 35 (12.7) |
| 2 | 73 (26.4) |
| 3 | 76 (27.5) |
| 4 | 36 (13.0) |
| 5 or more | 56 (20.3) |
| Number of households on Plot (mean (SD)) | 7.0 (5.0) |
| Electricity (n (%)) |  |
| Household meter connection | 111 (39.5) |
| Shared meter connection | 169 (60.1) |
| No connection | 1 (0.4) |
| Water Source (n (%)) |  |
| In own yard/plot | 74 (26.3) |
| In own dwelling | 18 (6.4) |
| Elsewhere | 189 (67.3) |

A majority (70%) of customers used public toilets before beginning CTG service. However, some were using a neighbour’s toilet (14%), or even a toilet already in their own compound (13%) or dwelling (2%). Public toilets were pay-per-use, with the most common fees being 0.50 GHS (US$0.09) (70%) or 0.40 GHS (US$0.07) (13%) per use. Structural quality of pre- CTG toilets, including the presence of solid walls and roofs without holes, the presence of locks, slab materials, and odour-reducing technologies, were generally reported to be good for both public and all other kinds of toilets. Most public toilets were flush or pour-flush toilets to septic tanks (79%), while non-public toilets were usually flush or pour-flush to septic tanks (30%), to pit latrines (27%), or VIP latrines (39%). Most public toilets were thought to be cleaned more than once a day (78%) and non-public toilets cleaned once a day (63%). Cleanliness was also generally good, with slabs (80%/79%) and pans (78%/77%) appearing clean at baseline when observed by enumerators. However, while 52% of public toilets had a place for handwashing with soap and water, only 16% of other toilets did. Smell reduction systems were often in place, including water seals (84%/21%), windows or vents (87%/59%), and ventilation pipes (90%/86%). Mean round-trip travel time to access public toilets including queuing and use was 14.3 minutes.

The living conditions of CTG customers were profiled using proxies for tenure; housing stock; infrastructure and resident’s perception of their neighbourhood, as per the Living Conditions Diamond analytical model, where 100% denotes an ideal living condition (Gulyani & Bassett, 2010; Scott et al., 2015). 99% of endline respondents lived in housing stock made from permanent building materials and although there was almost an even mix of landlords and tenants, 99% of CTG customers had no fear of being removed in the next two years and had lived in their current dwelling for an average of 11.6 years. Residents perceived their neighbourhoods to be clean (89%); a good location in the city (95%) and safe (98%). The infrastructure axis on the LCD is a composite of electricity; water; drainage; streetlighting; solid waste management; roads and an improved private toilet at home. This proxy was varied across the respondent group. It is noteworthy that 100% of CTG customers surveyed had access to electricity whereas 60% of use a public standpipe as their main source of water for handwashing and cooking. 34% had a solid waste management service to their plot. Comparing pre and post intervention, using the CBS service improved the living conditions profile of CTG customers (see figure 1).

**Figure 1:**
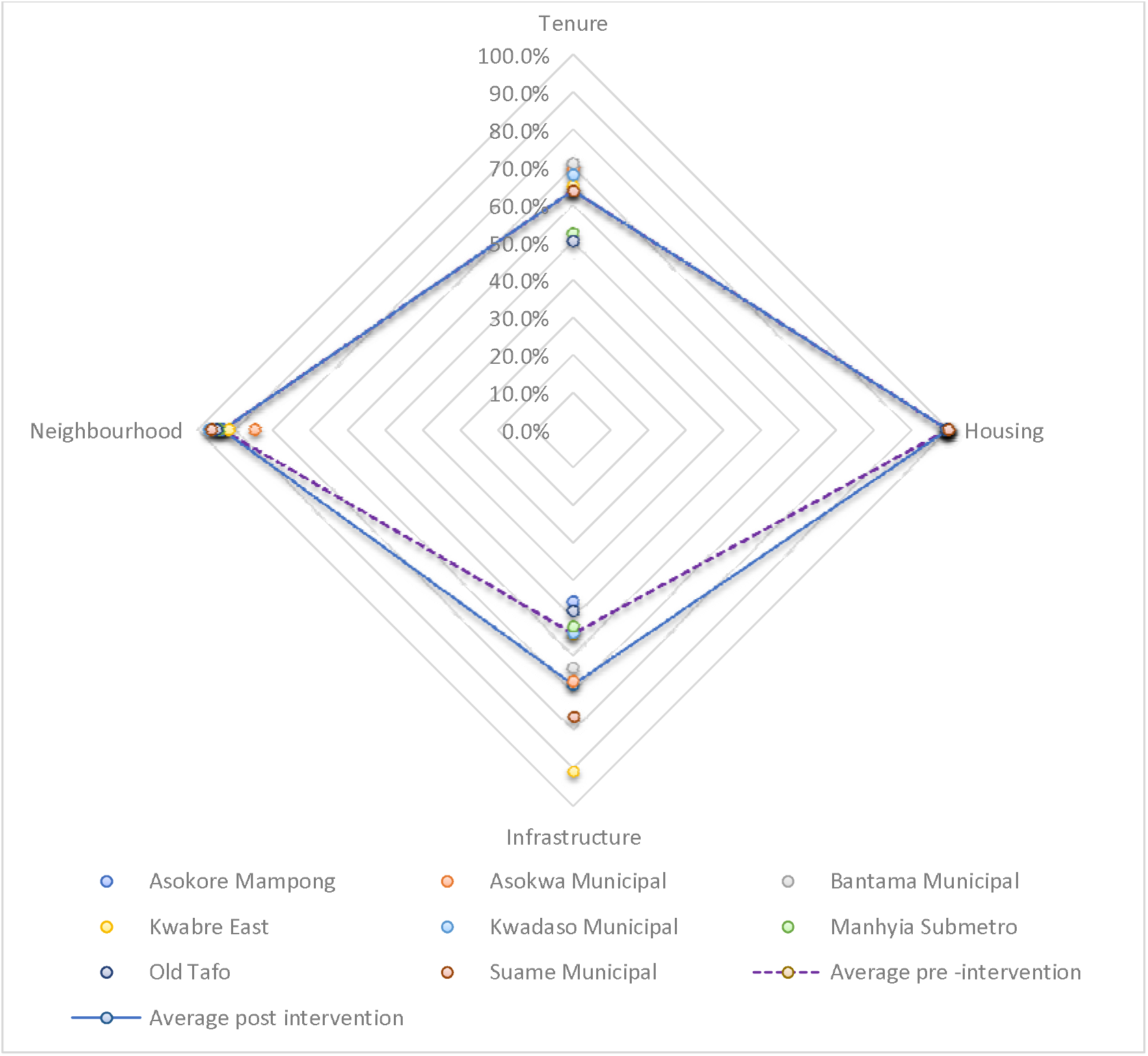
Living Conditions Diamond Values for CTG Customers.

### SERVICE QUALITY AND SATISFACTION

Overall, the CTG service functioned well across all of the measured indicators. Few customers experienced bad smells when the CTG toilet lid was closed (2%) or excessive smells when it was open (3%). Even fewer reported the toilet ever being too full to use (0.8%), and none said they had a problem with the toilet leaking. Only a small number (8%) had ever used a non-CTG toilet because of a problem with the CTG toilet. Almost all customers (96%) reported that their canister had been replaced twice a week as promised, though 11% reported having to wait for CTG staff three or more hours for replacements on average. However, it is not clear how substantially this impacts overall satisfaction with the service, as customers were sometimes around the home during these times anyway, and not missing work or other productive activities outside the home.

Sanitation satisfaction was scored on a scale from 0-4, with 0 representing very unsatisfied, 4 representing very satisfied, and 2 being neither satisfied nor unsatisfied (Table 3). The lowest mean pre-CTG score was for smell (1.57) and the highest for privacy (2.31). For those using CTG, scores across all categories increased by a minimum of 0.93 points to a maximum of 1.97 points on the four-point scale. Final satisfaction scores were all very high in an absolute sense, ranging from a low of 3.20 (cost) to a high of 3.56 (ease of access).

**Table 2:** Observed and Reported Baseline Toilet Characteristics

| Pre-CTG Toilet Characteristics | Value |
| --- | --- |
| Toilet location (%) |  |
| In own dwelling | 6 (2.2) |
| In own compound | 36 (13.3) |
| On neighbour's compound | 39 (14.4) |
| Public | 189 (70.0) |
| Sharing of Toilet with other households = Yes (%) | 251 (93.0) |
| Toilet Payment Type (%) |  |
| Per use | 186 (68.9) |
| None | 78 (28.9) |
| Other | 6 (2.2) |
| Fee per use (%) |  |
| 0.20 | 7 (3.8) |
| 0.30 | 16 (8.6) |
| 0.40 | 25 (13.4) |
| 0.45 | 1 (0.5) |
| 0.50 | 131 (70.4) |
| 0.60 | 3 (1.6) |
| 1.00 | 3 (1.6) |
| Typical number of daily uses (public toilets) (%) |  |
| 1 | 93 (48.4) |
| 2 | 98 (51.0) |
| 3+ | 1 (0.5) |
| Type of toilet (%) |  |
| Flush or pour-flush toilet to septic tank | 174 (64.4) |
| VIP latrine | 44 (16.3) |
| Flush or pour-flush toilet to pit latrine | 48 (17.8) |
| How often is toilet cleaned? (%) |  |
| More than once a day | 163 (60.4) |
| Once a day | 65 (24.1) |
| Several times a week | 38 (14.1) |
| Twice a week | 1 (0.4) |
| Don't Know | 3 (1.1) |
| Solid walls (made of durable materials without holes) (%) | 213 (99.1) |
| Solid roof (made of durable materials and without holes) (%) | 206 (95.8) |
| Solid door/locks (%) |  |
| Door locks inside and outside | 149 (69.3) |
| Door inside lock only | 37 (17.2) |
| Door outside lock only | 2 (0.9) |
| Door has no locks | 4 (1.9) |
| No door | 23 (10.7) |
| Slab cleanliness (%) |  |
| Appears clean | 168 (78.1) |
| Appears dirty or sandy/made of dirt | 44 (20.5) |
| Visible feces | 3 (1.4) |
| Pan material (%) |  |
| Ceramic | 115 (53.5) |
| Concrete | 98 (45.6) |
| Wood | 1 (0.5) |
| Dirt | 1 (0.5) |
| Pan cleanliness (%) |  |
| Appears clean | 164 (76.3) |
| Appears dirty or sandy/made of dirt | 37 (17.2) |
| Visible faeces | 14 (6.5) |
| Handwashing facility with soap/water (%) |  |
| Handwashing Soap and Water | 87 (40.5) |
| Handwashing Soap Only | 2 (0.9) |
| Handwashing Water Only | 30 (14.0) |
| Handwashing No Soap or Water | 1 (0.5) |
| No Handwashing Facility | 95 (44.2) |
| Water seal | 154 (71.6) |
| Simple cover | 26 (12.1) |
| Window or vent | 180 (83.7) |
| Ventilation pipe | 189 (87.9) |

**Table 3:** Customer Satisfaction and Quality of Life scores for Pre-CTG AND CTG Sanitation

| Satisfaction category | Pre-CTG | CTG | Diff | p-value |
| --- | --- | --- | --- | --- |
| Combine household duties and using toilet (%) | 1.59 (0.99) | 3.56 (0.53) | 1.97 | <0.001 |
| Avoiding unpleasant odour (%) | 1.57 (1.15) | 3.35 (0.75) | 1.78 | <0.001 |
| Use of toilet during menses (%) | 1.73 (0.94) | 3.40 (0.59) | 1.67 | <0.001 |
| Toilet comfort (%) | 1.95 (1.04) | 3.50 (0.55) | 1.55 | <0.001 |
| Ease of use (%) | 2.03 (0.94) | 3.52 (0.51) | 1.49 | <0.001 |
| Ease of access (%) | 2.11 (0.93) | 3.56 (0.52) | 1.45 | <0.001 |
| Toilet cleanliness (%) | 2.20 (0.93) | 3.52 (0.55) | 1.32 | <0.001 |
| Privacy of toilet use (%) | 2.31 (1.05) | 3.51 (0.58) | 1.20 | <0.001 |
| Cost of toilet usage (%) | 2.27 (1.01) | 3.20 (0.81) | 0.93 | <0.001 |
| SanQoL attributes: I can use the toilet... | Pre-CTG Mean | CTG Mean | Diff Mean | p-value |
| Without feeling disgusted | 1.86 (0.84) | 2.72 (0.45) | 0.86 | <0.001 |
| Without worrying about disease | 1.71 (0.88) | 2.70 (0.47) | 0.99 | <0.001 |
| In private, without being seen | 1.97 (1.06) | 2.74 (0.44) | 0.77 | <0.001 |
| Without feeling ashamed | 1.65 (1.05) | 2.76 (0.43) | 1.11 | <0.001 |
| With safety | 1.89 (1.05) | 2.75 (0.43) | 0.86 | <0.001 |

Sanitation-related Quality of Life (SanQoL) scores, measured on a 0 (Never able) to 3 (Always able) scale saw statistically significant increases across all five attributes. The largest single improvement was for shame (which had the lowest mean baseline level) and the smallest was for privacy (which was highest at baseline). Mean scores at endline were close to the maximum across all five attributes, and 66% of respondents reported the maximum possible score for all five attributes.

### GENDER

For the two explicitly gender-related variables, use of the CTG service led to women being satisfied or very satisfied with both their ability to practice MHM (97% compared to 23% pre-CTG) and to balance household duties and toilet use (98% compared to 18% pre-CTG). For other satisfaction categories, pre-CTG scores were lower for women than men in our sample at baseline, but increases in for ease of access, ease of use, and toilet cleanliness were significantly larger for women (Table 4). So, across all categories of sanitation satisfaction we measured, where women were worse off than men at baseline, the CTG service closed that gender gap entirely, and no statistically significant differences were observed between men and women using the CTG service. The same findings held for SanQoL, with lower SanQoL attribute values for women than men in our sample, all gender gaps closed, and a statistically significantly larger increase for safety.

**Table 4:** Satisfaction and SanQoL by Sanitation Service by Gender

| Satisfaction Category | Pre-CTG |  | CTG |  | p-value (DiD) |
| --- | --- | --- | --- | --- | --- |
|  | Female Mean (SD) | Male Mean (SD) | Female Mean (SD) | Male Mean (SD) |  |
| Avoiding unpleasant odour (%) | 1.54 (1.14) | 1.67 (1.22) | 3.37 (0.73) | 3.29 (0.80) | .285 |
| Avoiding unpleasant odour when not in use (%) | NA | NA | 3.47 (0.61) | 3.38 (0.69) | .311 |
| Combine household duties and using toilet (%) | 1.59 (0.99) | NA | 3.56 (0.53) | NA | <.001 |
| Use toilet during menses (%) | 1.73 (0.94) | NA | 3.40 (0.59) | NA | <.001 |
| Cost of toilet usage (%) | 2.23 (1.01) | 2.41 (1.02) | 3.21 (0.81) | 3.17 (0.81) | .246 |
| Ease of access (%) | 2.02 (0.92)* | 2.41 (0.90) | 3.58 (0.52) | 3.53 (0.53) | .003 |
| Ease of use (%) | 1.95 (0.94)* | 2.33 (0.90) | 3.54 (0.56) | 3.47 (0.53) | .003 |
| Privacy of toilet use (%) | 2.26 (1.04)* | 2.51 (1.04) | 3.53 (0.58) | 3.46 (0.58) | .058 |
| Toilet cleanliness (%) | 2.15 (0.94)* | 2.38 (0.90) | 3.54 (0.51) | 3.46 (0.53) | .041 |
| Toilet comfort (%) | 1.91 (1.03) | 2.07 (1.11) | 3.52 (0.55) | 3.43 (0.55) | .145 |
| SanQoL attributes: I can use the toilet... | Pre-CTG |  | CTG |  | p-value (DiD) |
|  | Female Mean (SD) | Male Mean (SD) | Female Mean (SD) | Male Mean (SD) |  |
| Without feeling disgusted | 1.81 (0.82) | 1.91 (0.90) | 2.75 (0.43) | 2.66 (0.48) | .142 |
| Without worrying about disease | 1.65 (0.87) | 1.73 (0.93) | 2.73 (0.46) | 2.63 (0.49) | .199 |
| In private, without being seen | 1.91 (1.06) | 2.06 (1.04) | 2.78 (0.42) | 2.67 (0.47) | .107 |
| Without feeling ashamed | 1.60 (1.04) | 1.70 (1.09) | 2.80 (0.40) | 2.67 (0.47) | .146 |
| With safety | 1.81 (1.07) | 2.09 (0.93) | 2.79 (0.41) | 2.68 (0.47) | .014 |

Note: \*: Statistically significant difference between Females and Males pre-CTG at $p < .05$ level

Although only a small number of customers had a household member excluded from using the toilet the family used pre-CTG due to physical or social reasons (5.2%), CTG service reduced this to only one of the 292 CTG customers reporting such a challenge (0.4%).

## Discussion

Overall, customers found that the Clean Team service met their needs and were satisfied with the service provided. While the cost of the CTG service was lower than the mean expenditure on public toilet fees, there were many customers who were paying both far more and far less than the monthly price of the CTG service beforehand. This suggests that while some may benefit primarily from financial advantages, others certainly prefer it for non-financial reasons. Objective indicators of sanitation quality for pre-CTG toilet structures were relatively high (e.g. materials used in the superstructure and containment type), though a lack of cleanliness and place for handwashing were sometimes a concern. CTG service quality was generally good, both as reported by customers and through enumerator observations of their CTG toilets. Few customers reported issues with toilets filling, smelling, leaking, or being replaced as scheduled. Therefore, the differences observed between pre- CTG and CTG service were a good estimate of how experiences might differ between relatively high-quality public toilets versus a well-run CBS service in general.

Satisfaction with the CTG service was high across all categories measured and was substantially higher than pre-CTG sanitation (except in cases where existing satisfaction had been high already). This suggests that although CBS is not necessarily the long-term aspiration for households, CBS provides a very positive user experience, rather than just being a small step up from public toilets—especially in dense urban settlements where permanent, private household toilets are not feasible. Baseline satisfaction and quality of life gaps between women and men were all closed. Considering explicitly gendered outcomes, almost all female customers were satisfied or very satisfied with CTG service compared to low rates of satisfaction pre-CTG. The number of people experiencing difficulty using the toilet also decreased drastically. This indicates that for those choosing the service, inequities due to gender or physical ability are all but eliminated.

One area for further investigation was satisfaction with the cost of the service. While CTG service was cheaper than pre-CTG sanitation options for a majority of customers, and satisfaction with costs increased from 47% to 85%, there was also more worry associated with paying for those costs, with those sometimes or always worrying about the cost of sanitation increasing from 19% to 31%. We hypothesize that this is because of the need to pay for CTG in a lump sum rather than the per-use payment for public toilets, and therefore the perceived risk of losing service more likely to lead a customer to worry. Financial products that allow customers to spread out their payments may be able to alleviate these concerns.

Understanding how CTG customers access other basic services (i.e. water, solid waste management) and customer development priorities may provide insights around opportunities for scaling up the service. First, employment was the second most commonly cited development priority of CTG customers, after roads, so CTG could consider promote their service as providing local employment opportunities. Second, only 34% of CTG customers had a door to door solid waste collection service. There may be potential efficiency gains worth exploring with solid waste management, which was cited as the third most common development priority.

These findings and the acknowledgement by the Joint Monitoring Programme (JMP) that CBS comprises a safely managed sanitation service (WHO/UNICEF, 2018) imply that policy and regulatory amendments to support CBS should be considered. The Ministry of Sanitation and Water Resources should consider a formal approval of the container-based sanitation as an approved sanitation technology option for Ghana. This should cover regulatory amendments in the National Building Code regarding CBS for domestic sanitation. Additionally, Metropolitan, Municipal and District Assemblies (MMDAs) should provide regulations for households and CBS service providers to ensure compliance with public health standards, covering: (1) safe disposal of anal cleansing materials, urine, grey water, (2) timeliness and quality of container emptying, (3) ensuring that the CBS technology and associated user behaviors do not result in unacceptably high fecal contamination exposures, and (4) reuse of treated faeces in agriculture.

There were several limitations to our study. First, due to funding constraints, we surveyed only customers and not a representative sample of the population. It is difficult to say how customers differed from the general population and therefore what the potential market size is for CBS. Second, while careful attention was given to using measures of satisfaction and quality of life that were as rigorous as possible, there is always the possibility of reporting bias or social desirability bias with psychometric questions. We made clear that we were independent of implementers, but respondents may have perceived differently. However, given the study objectives were to understand customer perceptions, not how the general population would perceive the service, these limitations minimally affect our key findings and recommendations in this study.

We suggest the following areas of future research on CBS based on the results of this study:

⍰ A survey of either non-buyers or the general population of CBS service areas to understand reasons for non-purchase of the service to better establish the market opportunity and service improvements that are desired by those not enrolling in the service
⍰ Understanding the costs and demand for CBS service improvements, including increased willingness to pay, to maximize the potential market for CBS
⍰ Adaptation of the service model to other settings, such as schools or shared plot toilets, where shared maintenance challenges need to be overcome, but may increase revenue opportunities for CTG
⍰ Understanding the relationship between demand for CBS and other basic services, specifically water and solid waste management.

## Conclusion

The evaluation found that CTG delivers a high-quality service that has significant impacts on the quality of life on CTG customers, as well as saving them money, reducing gender gaps in quality of life, and addressing the needs of those with physical disabilities and improving their living conditions.

Sanitation systems are a core element of public health infrastructure for cities. Ensuring equitable access to safely managed sanitation for all urban residents will likely require a range of pragmatic solutions. Container-based sanitation, if properly managed, is one such option, which ensures full containment of faecal waste. While one product or service does not fit the needs of all customers, and CBS service providers may consider other ways to expand their customer base through additional product and service offerings in the future, the current service plays a valuable role in achieving Citywide Inclusive Sanitation.

**Supplementary table 1:** REASONS WHY CUSTOMERS STOPPED USING CTG TOILET FACILITY AFTER INSTALLATION

| **Reasons for the dropouts** | **Counts (%)** |
| --- | --- |
| Space constraints | 25 (37%) |
| Customer relocated | 8 (12%) |
| Didn't understand payment policy/complain it's expensive | 8 (12%) |
| Household head/partner disagreed to use | 6 (9%) |
| Complaints of odour issues | 3 (4%) |
| Only once per week toilet pick | 3 (4%) |
| Landlord disallowed usage | 2 (3%) |
| Refused to give any reason | 2 (3%) |
| Other reasons | 10 (15%) |

## Data Availability

Data is available upon request and will be made publicly available upon publication.

